# Variation in Anticoagulation Practice for Atrial High-Rate Episodes: a Nationwide Cross-sectional Survey

**DOI:** 10.64898/2026.05.17.26353433

**Authors:** Kyi Zin Thant, Ibrahim Antoun, Kaung Myat Thu, Riyaz Somani, Zakariyya Vali, G Andre Ng, Mokhtar Ibrahim

## Abstract

**Background:** Atrial high-rate episodes (AHRE) detected by cardiac implantable electronic devices (CIEDs) are associated with increased thromboembolic risk, yet their clinical significance and optimal anticoagulation strategy remain uncertain, particularly in the absence of electrocardiogram (ECG)-confirmed atrial fibrillation.

**Methods:** We conducted a nationwide cross-sectional survey of UK clinicians involved in CIED follow-up. The survey assessed anticoagulation decision-making in AHRE, including episode-duration thresholds, cumulative burden, CHA_2_DS_2_-VA use, additional ECG monitoring, and anticoagulant choice. Only responses from UK-based consultant clinicians were included and analysed descriptively.

**Results:** A total of 51 responses were received; 38 met the inclusion criteria and were analysed. Most respondents (86.8%) reported having reviewed AHRE alerts within the preceding six months, indicating that AHRE are commonly encountered in clinical practice. A ≥24-hour episode was the most common threshold for anticoagulation (44.7%), although many clinicians reported lower thresholds or individualised approaches. Nearly half (44.7%) did not consider cumulative AHRE burden in decision-making. CHA_2_DS_2_-VA thresholds also varied, most commonly ≥2 or ≥1. Additional ECG monitoring was infrequently performed. Direct oral anticoagulants were universally preferred, with apixaban the most commonly selected agent (73.7%).

**Conclusion:** There is substantial variation in UK clinical practice regarding anticoagulation for AHRE, reflecting ongoing uncertainty and lack of clear guidance. These findings highlight the need for evidence-based thresholds to support consistent and informed clinical decision-making.

## Introduction

Atrial high-rate episodes (AHRE), detected through cardiac implantable electronic devices (CIEDs), are increasingly encountered in routine clinical practice. These subclinical atrial tachyarrhythmias have been associated with an increased risk of atrial fibrillation (AF) and thromboembolic events, yet their clinical significance, particularly in the absence of surface electrocardiogram (ECG)-confirmed AF, remains uncertain^1–3^. Earlier observational studies, including MOST and ASSERT, demonstrated associations between device-detected atrial tachyarrhythmias and increased risks of stroke, systemic embolism and progression to clinical AF.^4,5^ However, the absolute thromboembolic risk associated with shorter-duration AHRE remains incompletely defined, and uncertainty persists regarding the threshold at which anticoagulation may provide net clinical benefit.

Recent randomised trials have produced conflicting evidence regarding the role of anticoagulation in AHRE. The NOAH-AFNET 6 trial demonstrated no net clinical benefit of anticoagulation, largely due to increased bleeding risk, whereas the ARTESiA trial reported a reduction in stroke risk at the expense of higher bleeding rates.^6,7^ This discordance has contributed to uncertainty in clinical decision-making and highlights the lack of consensus regarding optimal management strategies.

Together, these findings highlight the uncertainty surrounding AHRE management and provide the rationale for examining real-world clinical practice.

Current European Society of Cardiology (ESC) guidelines acknowledge the uncertainty surrounding AHRE and do not provide definitive thresholds for anticoagulation; instead, they recommend individualised decision-making based on episode duration and patient risk profile^3^. In the absence of clear guidance, clinicians may adopt heterogeneous approaches influenced by personal experience, perceived risk, and extrapolation from clinical AF management.

This study aims to evaluate real-world clinical practice across the UK, focusing on anticoagulation thresholds, the use of CHA_2_DS_2_-VA scoring, and the role of additional monitoring in patients with AHRE.

## Methods

A cross-sectional survey was conducted to evaluate anticoagulation decision-making regarding atrial high-rate episodes (AHRE) among clinicians involved in the follow-up and management of patients with cardiac implantable electronic devices (CIEDs) in the United Kingdom. The survey was distributed electronically through professional networks, including direct email to colleagues, and was subsequently circulated more widely through the British Heart Rhythm Society (BHRS) mailing list using a convenience sampling approach. The survey was conducted between 29 October 2025 and 16 January 2026.

The survey was designed by the study team to assess key aspects of AHRE-related anticoagulation practice, including episode-duration thresholds, cumulative burden, use of CHA_2_DS_2_-VA scoring, ECG monitoring, and anticoagulant choice.

Because the survey distribution was broad, responses were screened for eligibility prior to analysis. Only responses from UK-based consultant clinicians were included in the final analysis. Responses from clinicians practising outside the UK and from non-consultant respondents were excluded. Eligibility was determined by manual review of self-reported respondent characteristics, including professional role and country of practice. Including only UK consultant clinicians in the final analysis allowed for greater consistency and relevance to UK clinical practice, although this may limit the generalisability of the findings to other healthcare settings.

Data were analysed descriptively and are presented as frequencies and percentages. Given the exploratory nature of this study, a formal sample size calculation was not performed. The study was designed to provide a descriptive overview of current practice patterns rather than to test a specific hypothesis. Participation was voluntary, and responses were anonymised prior to analysis. Formal research ethics committee approval was not required, as this study was conducted as a survey of current clinical practice.

## Results

A total of 51 survey responses were received. Following screening for eligibility, responses from clinicians practising outside the United Kingdom and those not at consultant level were excluded. After application of these criteria, 38 responses were included in the final analysis (Figure 1). Most respondents were cardiologists with a subspecialty focus in electrophysiology or cardiac devices (28/38, 73.7%), with the remainder from other cardiology subspecialties (10/38, 26.3%). Most respondents (86.8%) reported reviewing AHRE alerts within the preceding six months, indicating that AHRE are commonly encountered in clinical practice.

**Figure 1.**
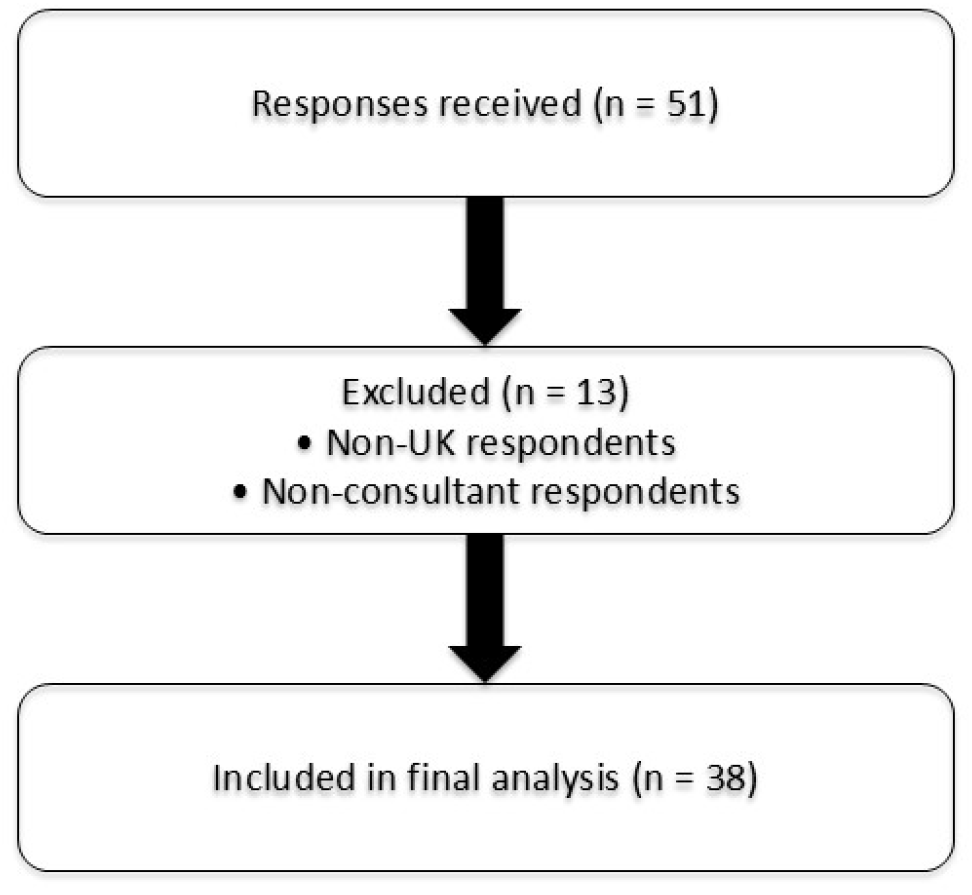
Flow diagram of respondent inclusion. Of 51 responses, 13 were excluded (non-UK and non-consultant respondents), leaving 38 included in the final analysis.

There was marked heterogeneity in thresholds for initiating anticoagulation. While a single-episode duration of ≥24 hours was the most commonly selected criterion (44.7%), over half of respondents reported using lower thresholds or individualised approaches. Approaches to cumulative AHRE burden also differed: 44.7% of respondents did not incorporate cumulative burden into decision-making, while others applied thresholds such as ≥5.5 hours or ≥24 hours. CHA_2_DS_2_-VA thresholds also differed, with most respondents adopting thresholds of ≥2 or ≥1.

The use of additional ECG monitoring prior to initiating anticoagulation was inconsistent, with most respondents reporting that they rarely or never performed further monitoring. Direct oral anticoagulants were universally preferred, with apixaban being the most commonly selected agent (73.7%). Detailed responses are summarised in Tables 1 and 2.

**Table 1.**
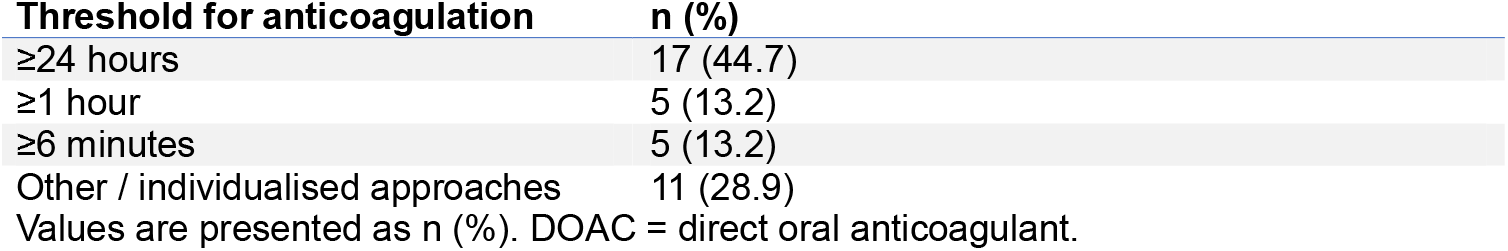
Reported thresholds for initiating anticoagulation among respondents.

| Threshold for anticoagulation | n (%) |
| --- | --- |
| ≥24 hours | 17 (44.7) |
| ≥1 hour | 5 (13.2) |
| ≥6 minutes | 5 (13.2) |
| Other / individualised approaches | 11 (28.9) |

Values are presented as n (%). DOAC = direct oral anticoagulant.

**Table 2.**
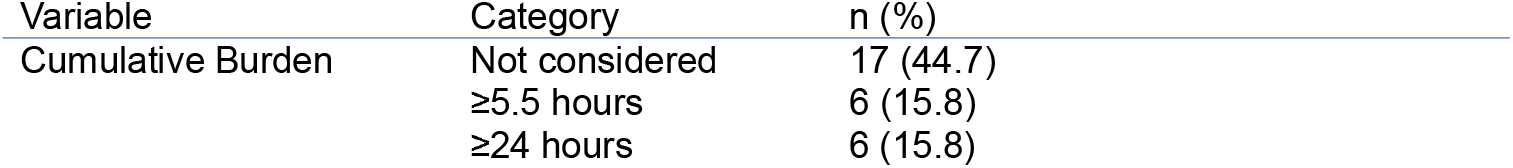
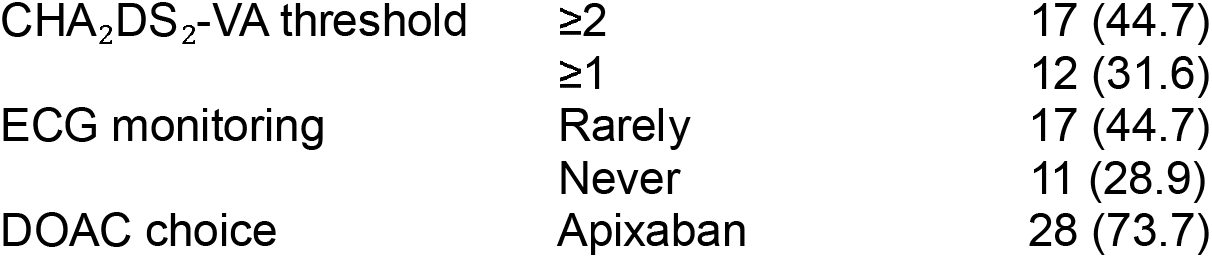
Clinician-reported factors and practices influencing anticoagulation decisions.

| Variable | Category | n (%) |
| --- | --- | --- |
| Cumulative Burden | Not considered | 17 (44.7) |
|  | ≥5.5 hours | 6 (15.8) |
|  | ≥24 hours | 6 (15.8) |
| CHA<sub>2</sub>DS<sub>2</sub>-VA threshold | ≥2 | 17 (44.7) |
|  | ≥1 | 12 (31.6) |
| ECG monitoring | Rarely | 17 (44.7) |
|  | Never | 11 (28.9) |
| DOAC choice | Apixaban | 28 (73.7) |

## Discussion

This survey demonstrates substantial heterogeneity in UK clinical practice regarding anticoagulation for AHRE, particularly in treatment initiation thresholds and the use of cumulative burden. These findings primarily reflect variation in clinician opinion and contemporary practice rather than evidence supporting any single management strategy. While nearly half of respondents used a ≥24-hour threshold for initiating anticoagulation, a significant proportion adopted lower thresholds or individualised approaches that incorporated the CHA_2_DS_2_-VA score and AHRE burden. This heterogeneity is unsurprising in light of evolving evidence regarding AHRE management. Recent ESC guidelines acknowledge the uncertainty surrounding anticoagulation in AHRE and recommend case-by-case clinical assessment based on episode duration and thromboembolic risk profile, without defining clear thresholds for treatment.^3^ Similarly, expert consensus statements and international guidelines continue to acknowledge the limited evidence base supporting anticoagulation in AHRE and emphasise the importance of individualised clinical decision-making in the absence of standardised treatment thresholds.^3,8^

Emerging evidence suggests that increasing AHRE burden is associated with progression to clinical atrial fibrillation and increased thromboembolic risk.^1^ Contemporary Europace literature has further highlighted the heterogeneous clinical profile of patients with AHRE detected through implantable devices, many of whom have substantial cardiovascular comorbidity and elevated baseline thromboembolic risk, further complicating anticoagulation decision-making.^9^ Multiple observational studies and meta-analyses have consistently demonstrated an association between device-detected atrial arrhythmias and increased thromboembolic risk, although the absolute risk appears lower than that observed in clinically diagnosed atrial fibrillation.^10–13^ Observational studies have also suggested a relationship between longer AHRE duration and higher thromboembolic risk, although proposed thresholds have varied substantially across studies.^5,8^ Proposed duration thresholds have ranged from episodes lasting several minutes to those exceeding 24 hours, contributing to variability in clinical practice and uncertainty regarding anticoagulation initiation.^11,14–16^

Recent randomised trials have produced conflicting findings. The ARTESiA trial demonstrated that apixaban reduced stroke and systemic embolism in patients with device-detected subclinical atrial fibrillation, although this was accompanied by increased major bleeding.^6^ In contrast, NOAH-AFNET 6 did not demonstrate a net clinical benefit of anticoagulation with edoxaban and reported higher bleeding rates.^7^ Together, these findings highlight the continuing ambiguity regarding which patients with AHRE may derive net benefit from anticoagulation and likely contribute to the observed variation in contemporary clinical practice. These findings are consistent with previous reviews and observational studies highlighting substantial uncertainty regarding optimal AHRE management in the absence of definitive evidence-based thresholds.^12,17^

Recent reviews have emphasised that uncertainty persists not only regarding stroke prevention strategies, but also regarding the broader prognostic implications of AHRE, including cardiovascular mortality and progression to clinical atrial fibrillation.^18^ Several factors likely contribute to this observed variation in practice. Firstly, AHRE represents a diagnostically ambiguous entity, as device-detected atrial tachyarrhythmias do not necessarily equate to clinically confirmed atrial fibrillation. Secondly, clinicians must balance competing risks, particularly the potential reduction in thromboembolism against the increased bleeding risk observed in recent trials. Thirdly, the absence of validated thresholds for AHRE duration or burden introduces uncertainty, leading clinicians to extrapolate from established atrial fibrillation management strategies. Finally, behavioural factors such as individual risk tolerance and medico-legal considerations may further influence decision-making.

Notably, nearly half of clinicians did not incorporate cumulative AHRE burden into decision-making. Given emerging evidence suggesting that total arrhythmia burden may be associated with thromboembolic risk^1^, this finding highlights a potential gap between evolving evidence and real-world practice, suggesting that cumulative AHRE burden is not yet routinely incorporated into clinical decision-making. The variability in CHA_2_DS_2_-VA thresholds further underscores the absence of consensus. Although most clinicians use established stroke risk scoring systems, the lack of standardised thresholds in AHRE contributes to inconsistent practice.

The limited use of additional ECG monitoring prior to anticoagulation may reflect several factors. While some clinicians may consider device-detected episodes sufficient to guide management, this finding may also reflect pragmatic constraints such as limited access to ambulatory monitoring or time pressures within clinical workflows. It may also reflect uncertainty regarding the necessity of ECG confirmation in the context of AHRE, given the lack of clear guideline recommendations. Previous studies have also highlighted ongoing uncertainty regarding the role of ECG confirmation and monitoring strategies in patients with device-detected atrial arrhythmias.^19,20^ The strong preference for apixaban is consistent with contemporary anticoagulation practice, although its role in patients with isolated AHRE remains uncertain. Similarly, emerging data regarding antiarrhythmic strategies in AHRE remain limited and investigational, further contributing to heterogeneity in management approaches.^21^

The observed variation in practice has important clinical implications. Inconsistent thresholds for anticoagulation may result in both under-treatment, with missed opportunities for stroke prevention, and over-treatment, exposing patients to unnecessary bleeding risk. Standardisation of practice, supported by clearer evidence and guideline recommendations, may help to reduce unwarranted variation, improve patient outcomes and reduce the likelihood of medico-legal challenges which are often associated with variations in clinical practice.

## Limitations

This study has several limitations. The survey was distributed via professional networks and society mailing lists using a convenience sampling approach, which may introduce selection bias. Additionally, clinicians with a particular interest in AHRE may have been more likely to respond to the survey, introducing additional response bias. Although responses were screened for eligibility, including exclusion of non-UK and non-consultant respondents, this process relied on self-reported data and manual review, which may introduce misclassification bias.

The sample size was modest, and responses were self-reported, introducing potential response bias. Additionally, the study population was limited to UK consultant clinicians, which may affect generalisability to other geographies. Clinical outcomes were not assessed. Respondents were predominantly electrophysiologists or clinicians with a subspecialty interest in cardiac devices, which may limit the representativeness of the findings to broader general cardiology practice.

## Conclusion

There is substantial variation in UK practice regarding anticoagulation for AHRE, particularly in treatment initiation thresholds and the use of cumulative burden. This variation may have important consequences for patient care, with potential risks of both over- and under-anticoagulation. Clearer, evidence-based guidance is needed to support more consistent and informed clinical decision-making.

Further prospective studies are needed to clarify which patients with AHRE derive net benefit from anticoagulation, at what episode duration or cumulative burden treatment should be considered, and what role ECG confirmation should play in clinical decision-making.

## Data Availability

All data produced in the present study are available upon reasonable request to the authors

## Acknowledgments

The authors thank Dr. Myo Thiha Soe for substantial assistance with survey distribution.

